# A Retrospective Cohort Study of the 2018 Angiotensin Receptor Blocker (ARB) Recalls and Subsequent Drug Shortages on Patients with Hypertension

**DOI:** 10.1101/2023.08.17.23294250

**Authors:** Joshua W Devine, Mina Tadrous, Inmaculada Hernandez, Katherine Callaway Kim, Scott D Rothenberger, Nandita Mukhopadhyay, Walid F Gellad, Katie J Suda

## Abstract

**Purpose:** In July 2018, the Food and Drug Administration recalled valsartan due to carcinogenic impurities, leading to an unprecedented drug shortage leading to management challenges impacting a large population of valsartan users. However, the extent to which the valsartan recall impacted clinical outcomes is unknown. Our objective was to compare the risk of adverse events between patients with hypertension using valsartan and a propensity-score-matched group of patients using non-recalled angiotensin receptor blockers (ARBs) and angiotensin converting enzyme-inhibitors (ACE-Is).

**Methods:** We conducted a retrospective cohort analysis of Optum’s de-identified Clinformatics® Datamart (July 2017–January 2019). Adults with hypertension who received valsartan were compared to persons who received non-recalled ARBs and ACE-Is for 1 year prior to and on the recall date. The primary outcomes were measured in the 6 months following the recall date included: 1) a composite measure of all-cause hospitalization, all-cause emergency department (ED) and all-cause urgent care (UC) visit, 2) a composite cardiac event measure of hospitalizations for acute myocardial infarction and hospitalizations/ED/UC visits for stroke/transient ischemic attack, heart failure or hypertension. Cox proportional hazard models compared the risk of outcomes between propensity-score-matched treatment groups.

**Results:** Of adults with hypertension, 76,934 received valsartan at the time of the recall and 509,742 received a non-recalled ARB/ACE-I. Valsartan use at the time of the recall was associated with a combined increased risk of all-cause hospitalization, ED or UC use (HR 1.02; 95% CI: 1.00–1.04) and of the composite of cardiac events (HR 1.22; 95% CI: 1.15–1.29) within six-months after the recall.

**Conclusions:** The national valsartan recall and subsequent shortage had negative consequences for patients with hypertension. As the threat of shortages increases for drugs that treat common outpatient conditions, systems at the local- and national-levels need to be strengthened to protect patients from drug shortages by providing them with safe and reliable alternatives.

## INTRODUCTION

One of the largest ever retail drug shortages began in July 2018 when the Food and Drug Administration (FDA) recalled several generic versions of valsartan. Valsartan belongs to the angiotensin receptor blocker (ARB) medication class and is among low-cost, first-line treatments for hypertension and other cardiac-related conditions recommended by the American Heart Association (AHA) with the highest level of evidence.^1,2^ Access to this critical medication was threatened when FDA discovered carcinogenic impurities in the raw active ingredient.^3–5^ Although absolute cancer risk was low, the impurities resulted in recalls of over two dozen products, impacting millions of patients.^4,6^ Removal of contaminated valsartan from pharmacy shelves resulted in a global drug shortages, some lasting years.^7–9^

In the year of valsartan’s recall (2017-2018), 51% of American men and 40% of American women over 18 years old had hypertension.^10^ Hypertension is a major risk factor for cardiovascular disease, which is the leading cause of death for adults in the U.S. and globally.^11,12^ In addition to lifestyle interventions, pharmacologic treatment to lower blood pressure – including with valsartan and other renin-angiotensin-system (RAS)-acting agents - decreases the incidence of heart failure, stroke, and myocardial infarction.^1,2^ Approximately 80 million valsartan prescriptions were dispensed prior to the recall.^13^ The 2018 recalls disrupted nearly half of U.S. persons use of valsartan,^14–17^ with potentially catastrophic impacts on patients’ blood pressure control and downstream risk for serious adverse cardiovascular events.^4,5^ Previous, non-recall-related gaps in anti-hypertension treatment have been associated with increased all-cause and cardiovascular-disease-specific emergency department visits, hospitalizations, and mortality.^18–21^ Additionally, two studies from Alberta and Ontario, Canada found increases in hypertension-related emergency department visits among valsartan users immediately after Health Canada’s recall.^22,23^ However, the Canadian studies did not use a comparison group and the downstream clinical impact of the recall and subsequent valsartan shortage on U.S. patients with hypertension is unknown.

The primary objective of our study was to compare the cumulative incidence of major cardiovascular outcomes among U.S. adults with hypertension receiving valsartan at the time of the 2018 valsartan recall and subsequent drug shortage to those receiving similar drugs that were not affected by the recall. Our study adds to existing literature by comparing outcomes among impacted valsartan users versus users of other RAS-acting drugs which were not recalled – e.g., ACE-Inhibitors, or ACE-Is, and non-recalled ARBs. Our results provide evidence on the negative health impacts of drug recalls on a large population of patients with chronic cardiovascular disease which can motivate policies to improve quality in U.S. retail drug supply chains.

## METHODS

### Study Design and Data Source

We conducted a retrospective cohort study using Optum’s de-identified Clinformatics® Datamart Database, which includes medical claims, pharmacy claims, enrollment, and demographic information for patients with commercial or Medicare Advantage coverage from a large national health insurer. Our study period comprised one year prior to and 6 months after the date of the valsartan recall in July 2018 (July 2017 – January 2019). The University of Pittsburgh Institutional Review Board approved our study (#21060160). We followed STROBE reporting guidelines.

### Study Population

The study population included patients aged 18 years of age or older with hypertension who were active users (defined below) of either valsartan or a comparison non-recalled ARB/ACE-I on valsartan’s recall date (July 13 2018). We defined hypertension using a validated algorithm recommended by the Agency for Healthcare Research and Quality (AHRQ).^24^ Patients were included if they had any hospitalization (inpatient) or two ambulatory claims (outpatient medical/physician/advanced practice provider visit, ED visit, urgent care visit) with a hypertension diagnosis within two years. Patients were excluded if they had a diagnosis of heart failure, or were pregnant or delivered a baby in the previous 365 days.

The index date for our study was defined as the date that the FDA announced the valsartan recall (July 13, 2018). [7] Follow-up started on the day after the index date. Patients were included in the study cohort if they were continuous users of valsartan or a non-recalled ACE-I/ARB during the 1-year pre-index baseline period (July 13, 2017 – July 12, 2018) and current users on valsartan’s recall date (Appendix Table 1). We defined continuous use as having a medication possession ratio (MPR) of ≥80% pre-index.^25,26^ MPR was calculated as the sum of the days’ supply for all fills of a given drug, divided by the number of days in the time period (365 days). Users who switched between agents within the same class (e.g., ARB-to-ARB) were captured as continuous users.

### Patient Characteristics

Baseline patient demographic characteristics, prescription medication fills, and presence of comorbidities were collected during the pre-index date period. Demographic factors included sex, age, race, ethnicity, insurance status/policy, and geographic region. Comorbidities were operationalized using a composite Elixhauser comorbidity score based on ICD-10-CM diagnosis categories and the Swiss algorithm, and the comorbidity score was adjusted for in analyses.^27^

### Outcome Measures

The primary outcomes included 1) a composite measure for all-cause hospitalization, emergency department (ED) use, and urgent care (UC) use; 2) a composite measure of cardiac events, including hospitalization for myocardial infarction and hospitalization, ED/UC visits for stroke, transient ischemic attack (TIA), heart failure, or hypertension as the principal diagnosis.^28^ We also reported each component separately. We also defined several secondary outcomes including hospitalizations and ED/UC visits for hypertension, heart failure, and TIA/stroke. To avoid misclassification of acute myocardial infarction for other acute cardiac events, we limited this outcome to events coded during a hospitalization.^28,29^ All events were measured within the 6-month period following our index date (August 2018 – January 2019).

### Propensity Score Development

We employed propensity score-weighting, a quasi-experimental approach in the estimation of treatment effects, to minimize confounding and bias. First, the probability of being an exposed case or an unexposed control was modeled using logistic regression with all patient characteristics as predictors, and the estimated probabilities served as propensity scores. We then calculated inverse probability of treatment (IPT) weights for each patient by taking the inverse of their propensity scores, and IPT weighting (IPTW) was employed in all outcome analyses for the estimation of average treatment effects (ATEs). Specifically, stabilized IPTW-ATE weights were calculated and applied in regression modeling by adjusting the weights by the marginal probability of being a case or control. Balance in baseline characteristics between groups was assessed before and after weighting the sample with IPTW-ATE weights using standardized mean differences (SMDs), and adequate balance was defined as a SMD less than 0.10.

### Statistical Analysis

For all outcomes we evaluated whether the hazard (i.e., instantaneous risk) of adverse outcomes differed significantly between cases and controls using Cox proportional hazards (PH) regression models. Treatment group status served as the main predictor variable, and stabilized IPTW-ATE weights were incorporated in the fitting of models. The index date served as time zero, and patients whose insurance eligibility period expired before the six-month observation period without renewal were right-censored at the expiration date. We carried out all analyses before and after applying the weights, to provide unadjusted and adjusted results, respectively.

We estimated hazard ratios (HRs) associated with group status and corresponding 95% confidence intervals (CIs). Forest plots of hazard ratios and weighted Kaplan-Meier survival curves were constructed. Mortality was not included as an outcome because exact dates of death were not available in the dataset.

All p-values are two-sided, we set our Type 1 error rate at 0.05, and no adjustments for multiplicity were made. The R statistical language was used for all analyses; specifically, the core stats package, R WeightIt, and the Survival package.^30,31^ Analyses were conducted from August 2022 – July 2023.

## RESULTS

### Baseline Patient Characteristics

The study cohort consisted of 586,406 individuals with hypertension including 76,934 (13%) patients on valsartan when the FDA recall announcement occurred and 509,742 (87%) comparison individuals on ARBs or ACE-Is that were not subject to the FDA recall (Figure 1). The mean age of the study cohort was 69 years (SD 11.1), 49.6% were male, and 69.6% were white. The distributions of baseline characteristics are summarized in Table 1, stratified by exposure to the FDA recall at index date with standardized mean differences (SMDs) and weighted proportions before and after propensity-score weighting. Following propensity score weighting, baseline characteristics were well balanced between the exposed group and unexposed group with all SMDs ≤0.011 (Table 1; complete characteristics are included in Appendix Table 2).

**Table 1.** Comparison of valsartan users and other ARB/ACE-I users before and after propensity-score weighting for patient characteristics at the time of FDA recall announcement.

| Variable | Before Inverse Probability Weighting |  |  | After Inverse Probability Weighting |  |  |
| --- | --- | --- | --- | --- | --- | --- |
|  | *Valsartan Cohort (Exposed)* | *Other ACE-I/ARB Cohort (Unexposed)* | *SMD* | *Weighted Mean/Proportion (Exposed)* | *Weighted Mean/Proportion (Unexposed)* | *SMD* |
| N | 76,934 | 509,471 |  |  |  |  |
| Age – mean (SD) | 70.5 (10.8) | 69.3 (11.2) | **0.109** | 69.4 | 69.4 | <0.01 |
| Male sex – no. (%) | 31,720 (41.2) | 259,098 (50.9) | **0.194** | 0.496 | 0.498 | <0.01 |
| Race – no. (%) |  |  | **0.178** |  |  | 0.011 |
| White | 48,736 (63.3) | 359,684 (70.6) |  | 0.701 | 0.697 |  |
| Black | 12,280 (16.0) | 62,944 (12.4) |  | 0.128 | 0.128 |  |
| Hispanic | 9,910 (12.9) | 56,464 (11.1) |  | 0.110 | 0.113 |  |
| Asian | 3,482 (4.5) | 12,814 (2.5) |  | 0.027 | 0.028 |  |
| Unknown | 2,526 (3.3) | 17,566 (3.4) |  | 0.034 | 0.034 |  |
| Geographic Region – no. (%) |  |  | **0.247** |  |  | 0.011 |
| Midwest-East | 9,387 (12.2) | 77,510 (15.2) |  | 0.148 | 0.148 |  |
| Midwest-West | 2,813 (3.7) | 35,885 (7.0) |  | 0.066 | 0.066 |  |
| Northeast-Mid Atlantic | 8,591 (11.2) | 36,684 (7.2) |  | 0.076 | 0.077 |  |
| Northeast-New England | 2,416 (3.1) | 18,589 (3.6) |  | 0.036 | 0.036 |  |
| South-East South Central | 4,087 (5.3) | 26,526 (5.2) |  | 0.052 | 0.052 |  |
| South-South Atlantic | 24,880 (32.3) | 142,140 (27.9) |  | 0.283 | 0.285 |  |
| South-West South Central | 11,849 (15.4) | 72,870 (14.3) |  | 0.148 | 0.145 |  |
| West-Mountain | 4,785 (6.2) | 42,917 (8.4) |  | 0.081 | 0.081 |  |
| West-Pacific | 8,126 (10.6) | 56,351 (11.1) |  | 0.109 | 0.110 |  |
| Age Category – no. (%) |  |  | **0.1** |  |  | <0.01 |
| 18-64 | 20,543 (26.7) | 155,325 (30.5) |  | 0.303 | 0.300 |  |
| 65-74 | 30,247 (39.3) | 201,297 (39.5) |  | 0.395 | 0.395 |  |
| 75+ | 26,144 (34.0) | 152,850 (30.0) |  | 0.302 | 0.305 |  |
| History of MI – no. (%) | 296 (0.4) | 1,646 (0.3) | 0.01 | 0.003 | 0.003 | <0.01 |
| Elixhauser Comorbidity Score Pre-Index – mean (SD) | -0.53 (8.26) | -0.65 (7.97) | 0.015 | -0.6347 | -0.6468 | <0.01 |
| Elixhauser Comorbidity Binary Indicators – no. (%)* |  |  |  |  |  |  |
| Hypertension, complicated | 9,180 (11.9) | 48,906 (9.6) | 0.075 | 0.0990 | 0.0991 | <0.01 |
| Hypothyroidism | 13,854 (18.0) | 77,779 (15.3) | 0.074 | 0.1562 | 0.1558 | <0.01 |
| Valvular disease | 6,046 (7.9) | 31,220 (6.1) | 0.068 | 0.0635 | 0.0632 | <0.01 |
| Obesity | 14,840 (19.3) | 86,229 (16.9) | 0.061 | 0.1724 | 0.1723 | <0.01 |
| ARB/ACE-I Pre-Index Adherence Percentage – mean (SD) | 0.95 (0.05) | 0.95 (0.05) | 0.023 | 0.9467 | 0.9466 | <0.01 |

Abbreviations: SMD=standardized mean difference; ACE-Is=angiotensin-converting enzyme inhibitor; ARBs=angiotensin-receptor blocker; MI=myocardial infarction.
The propensity-score matching utilized the Elixhauser comorbidity score along with 31 binary disease specific predictors; the table above reported binary indicators that had SMD > 0.06; a full list is available in the online appendix; all SMD values > 0.1 are shown in bold.

**Figure 1.**
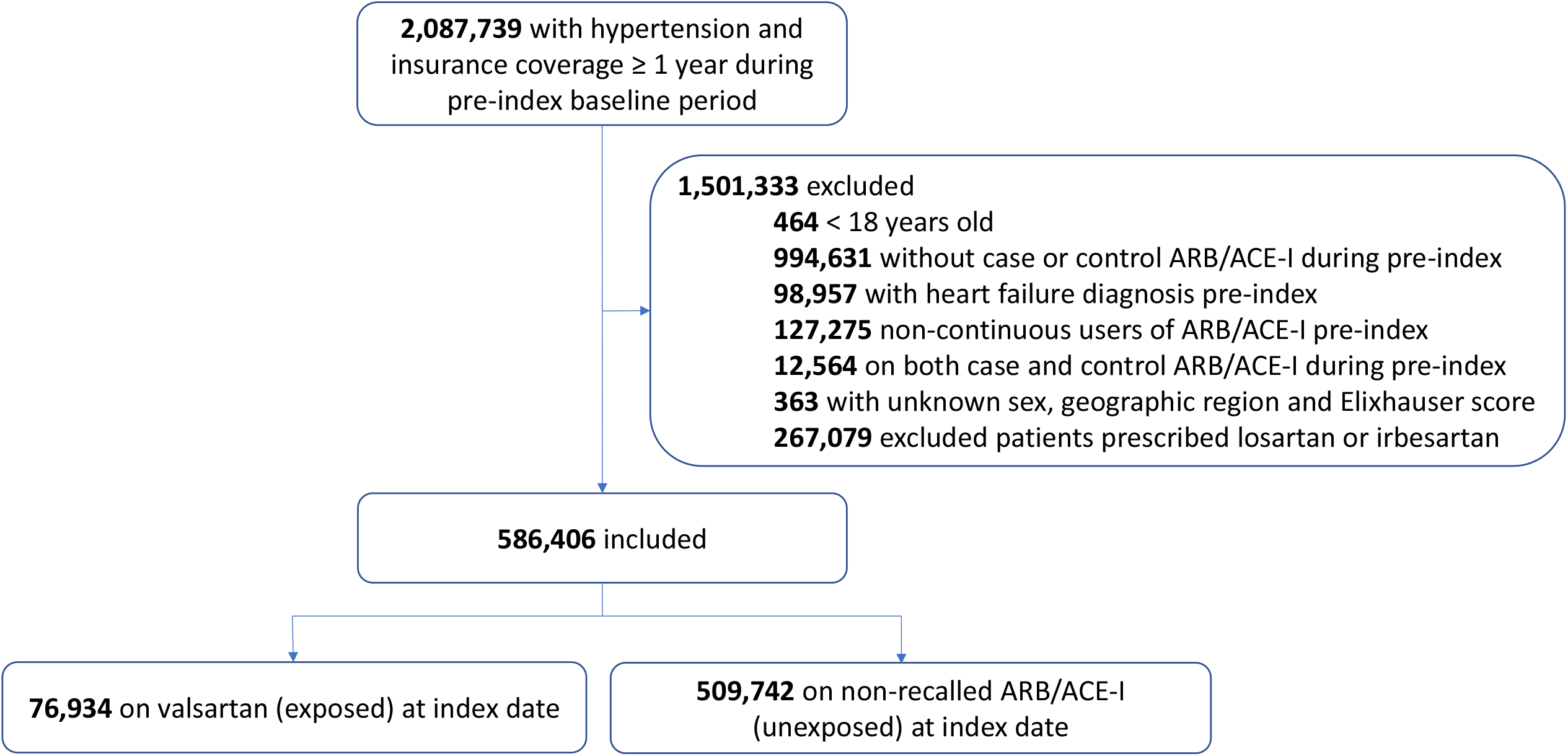
Cohort assignment for patients with hypertension on ARB/ACE-Is affected by a valsartan recall (exposed group) and those who did not experience a recall (unexposed group) ARB= angiotensin receptor blockers; ACE-Is= angiotensin-converting enzyme inhibitors CONSORT flow diagram to show construction of the study cohort and inclusion and exclusion criteria for the exposed group (valsartan) and unexposed group (non-recalled ARB and ACE-I). Index date was defined as the date the Food and Drug Administration (FDA) issued the valsartan recall (July 13, 2018).

### Primary All-Cause Study Outcomes

Nearly one-fifth of the cohort had at least one hospitalization, ED, or UC visit within 6-months following the FDA recall (19.2% [N=14,744] in exposed; 17.9% [91,378] in unexposed) (Table 2). All-cause hospitalization occurred at a similar frequency between groups (6.3% [N=4861] in valsartan users; 6.1% [N=31,185] among comparison drug users), with small increases in the valsartan group for ED visits (14.8% [N=11,376] in exposed; 13.9% [N=70,871] in unexposed), UC visits (3.5% [N=2712] in exposed; 3.1% [N=15,978] in unexposed), and cardiac events (2.1% [N=1607] in exposed; 1.7% [N=8799] in unexposed).

**Table 2.**
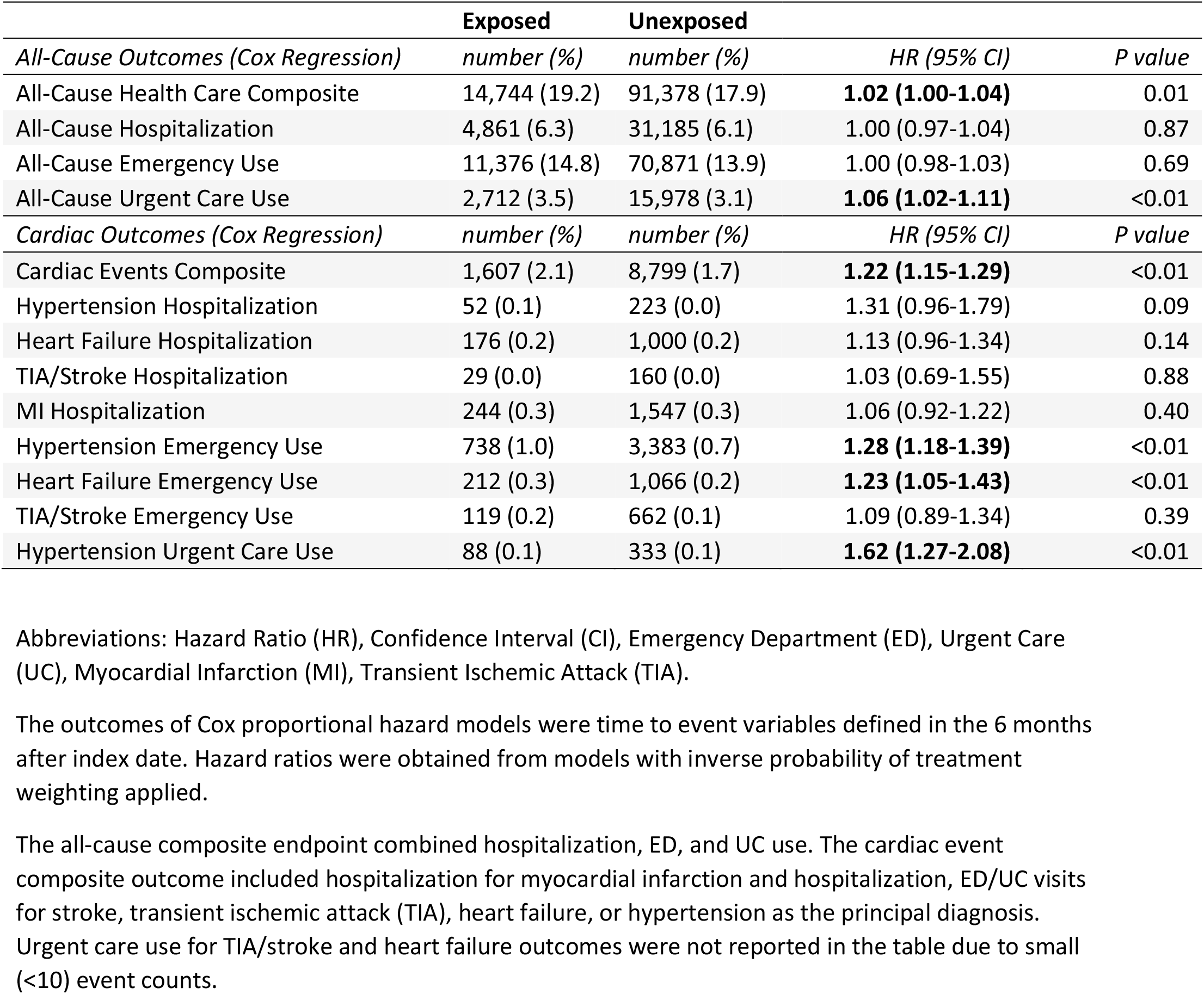
The impact of exposure to a valsartan drug recall on clinical outcomes in multivariable propensity-score weighted analysis.

|  | **Exposed** | **Unexposed** |  |  |
| --- | --- | --- | --- | --- |
| *All-Cause Outcomes (Cox Regression)* | *number (%)* | *number (%)* | *HR (95% CI)* | *P value* |
| All-Cause Health Care Composite | 14,744 (19.2) | 91,378 (17.9) | **1.02 (1.00-1.04)** | 0.01 |
| All-Cause Hospitalization | 4,861 (6.3) | 31,185 (6.1) | 1.00 (0.97-1.04) | 0.87 |
| All-Cause Emergency Use | 11,376 (14.8) | 70,871 (13.9) | 1.00 (0.98-1.03) | 0.69 |
| All-Cause Urgent Care Use | 2,712 (3.5) | 15,978 (3.1) | **1.06 (1.02-1.11)** | <0.01 |
| *Cardiac Outcomes (Cox Regression)* | *number (%)* | *number (%)* | *HR (95% CI)* | *P value* |
| Cardiac Events Composite | 1,607 (2.1) | 8,799 (1.7) | **1.22 (1.15-1.29)** | <0.01 |
| Hypertension Hospitalization | 52 (0.1) | 223 (0.0) | 1.31 (0.96-1.79) | 0.09 |
| Heart Failure Hospitalization | 176 (0.2) | 1,000 (0.2) | 1.13 (0.96-1.34) | 0.14 |
| TIA/Stroke Hospitalization | 29 (0.0) | 160 (0.0) | 1.03 (0.69-1.55) | 0.88 |
| MI Hospitalization | 244 (0.3) | 1,547 (0.3) | 1.06 (0.92-1.22) | 0.40 |
| Hypertension Emergency Use | 738 (1.0) | 3,383 (0.7) | **1.28 (1.18-1.39)** | <0.01 |
| Heart Failure Emergency Use | 212 (0.3) | 1,066 (0.2) | **1.23 (1.05-1.43)** | <0.01 |
| TIA/Stroke Emergency Use | 119 (0.2) | 662 (0.1) | 1.09 (0.89-1.34) | 0.39 |
| Hypertension Urgent Care Use | 88 (0.1) | 333 (0.1) | **1.62 (1.27-2.08)** | <0.01 |

Abbreviations: Hazard Ratio (HR), Confidence Interval (CI), Emergency Department (ED), Urgent Care (UC), Myocardial Infarction (MI), Transient Ischemic Attack (TIA).
The outcomes of Cox proportional hazard models were time to event variables defined in the 6 months after index date. Hazard ratios were obtained from models with inverse probability of treatment weighting applied.
The all-cause composite endpoint combined hospitalization, ED, and UC use. The cardiac event composite outcome included hospitalization for myocardial infarction and hospitalization, ED/UC visits for stroke, transient ischemic attack (TIA), heart failure, or hypertension as the principal diagnosis. Urgent care use for TIA/stroke and heart failure outcomes were not reported in the table due to small (<10) event counts.

After IPTW, exposure to the valsartan recall was associated with a combined increased risk of all-cause hospitalization, ED or UC use (HR 1.02; 95% CI 1.00 – 1.04). Individuals in the valsartan group had similar hazard of all-cause hospitalization (HR 1.00; 95% CI 0.97 – 1.04) and all-cause ED use (HR 1.00; 95% CI 0.98 – 1.03), but higher hazard of urgent care use (HR 1.06; 95% CI 1.02 – 1.11) six-months following the recall, compared to individuals taking non-recalled drugs (Table 2). The risk of cardiac events was also higher among valsartan users compared with individuals on non-recalled ARBs/ACE-Is (HR 1.22; 95% CI 1.15 – 1.29).

### Secondary Disease-Specific Clinical Outcomes

The results from our secondary analyses examining disease-specific outcomes are shown in Table 2. In the 6 months post-valsartan-recall, the hazards of hospitalization for hypertension (HR 1.31; 95% CI 0.96 – 1.79), heart failure (HR 1.13; 95% CI 0.96 – 1.34), TIA/stroke (HR 1.03; 95% CI 0.69 – 1.55) and MI (HR 1.06; 95% CI 0.92 – 1.22) did not differ between the groups. ED use for hypertension (HR 1.28; 95% CI 1.18 – 1.39) and heart failure (HR 1.23; 95% CI 1.05-1.43) were higher in the valsartan group while ED use for TIA/stroke did not differ between groups (HR 1.09; 95% CI 0.89 – 1.34). Hypertension-related urgent care use was also greater among the valsartan group compared with individuals on non-recalled ARBs/ACE-Is (HR 1.62; 95% CI 1.27-2.08).

## DISCUSSION

In a nationwide cohort of patients with hypertension, individuals taking valsartan at the time of the FDA recall had an increased risk of all-cause hospitalization, ED, or UC visit compared to users of non-recalled ARBs and ACE-Is. Importantly, valsartan users also had increased risk of cardiac events within 6-months after the recall. Therefore, our results demonstrate the effect that a recall and subsequent shortage of an oral agent used for a common chronic condition (i.e., hypertension) can have on health care utilization and patient outcomes.

Our study makes several important contributions to existing literature on the 2018 valsartan recall. Previous reports have been mixed on the extent the valsartan recall and subsequent shortage impacted patient outcomes. Two percent of Veterans receiving recalled valsartan had an ED visit with a hypertensive emergency within 3 months post-recall.^32^ Another Canadian study using an ecological design reported small increases in hypertension-related ED visits at the population-level in adults ≥65 years of age with temporal increases in all-cause ED visits and hospitalizations.^22^ However, attributing these events to the recall is difficult because neither study included a comparison group. Our study addressed this limitation and found increased cardiac events among valsartan users in the six months post-recall, compared to similar, unaffected patients who were using non-recalled ARBs/ACE-Is at baseline. As with our outcomes, it is unclear if the reason for increasing health care utilization was related to cardiac events or other diagnoses due to loss of blood press control or the protective effect of ARBs, patients seeking alternative medication, or adverse drug events associated with medication switching. Regardless of the reason, the use of the ED or UC due to a drug shortage is avoidable, especially for medication switches.

Our analyses identified an association of a drug recall followed by a shortage and poor patient outcomes in patients with hypertension, a condition affecting nearly half of the U.S. adult population.^33^ Patients may respond differently to alternative medications, including increased/decreased therapeutic affect (e.g., hypotension, untreated hypertension), abnormal laboratory values (e.g., hyperkalemia), adverse drug events, or time to establish blood pressure control.^4,34^ As seen in our study, retail drug shortages therefore have potential to increase patients’ risk for medication disruptions and subsequent adverse health events. Sequelae of the valsartan recall occurred despite having some non-recalled versions of valsartan and suitable alternatives available (non-recalled ARBs and ACE-Is) in the U.S. Thus, the impact of other outpatient drug shortages with few alternatives available may have an even larger impact on patient outcomes.

Besides our results demonstrating the downstream implications of a drug recall and subsequent shortage on patient outcomes, other investigators have reported disruption in patient’s medication regimens.^14,17,35^ A national study found that 49% of valsartan users switched to a non-valsartan ARB, ACE-I, or calcium channel blocker.^14^ However, a study of a single Veterans Affairs (VA) facility found that only 4% of Veterans on valsartan switched their medication to an alternative antihypertensive post-2018-recall.^32^ Unlike the national analysis across multiple-health care systems, the integrated VA health care system had the ability to identify and educate affected patients. A mailing included instructions for Veterans to continue taking their current valsartan supply, assurances that subsequent prescriptions would be filled with non-recalled valsartan, and education on the risk of uncontrolled blood pressure and potential for carcinogen exposure.^32^ The extent to which other health-systems or pharmacies were able to provide outreach to affected patients is unknown.

Our results are not without limitations. First, we used administrative claims data which can be impacted by confounding and measurement error. We attempted to control for this by using propensity score-weighting which performed well with standardized mean differences ≤0.011 between groups. We also used a comparison group of medications (ARBs/ACE-I) that are considered to have similar indications and effectiveness. Second, a 6-month time window may not be sufficient for patients to run out of residual valsartan supply and/or develop adverse cardiac outcomes, and longer term follow up is needed. Third, we did not assess blood pressure control or treatment discontinuations or switches because of inability to access detailed clinical data. Fourth, patient race and ethnicity variables are imputed by the data provider and not self-identified. Lastly, to examine the association between the drug recall with outcomes, we assessed an adherent cohort. While the thresholds we applied are recommended methods, adherent patients may have different medication use behaviors.^25,26^

This study has far-reaching implications. Shortages of non-injectables are becoming more common, now comprising over 50% of drug shortage reports annually.^36,37^ Substituting drugs on shortage for alternative medications may be more complicated in the fragmented outpatient health care system, compared to acute care facilities where the health care team have access to a single health record and pharmacy infrastructure. There are likely other implications including cost of additional health care resources required to prescribe an alternate medication, pharmacist time to communicate with prescribers and procure alternate medications, and increased medication costs.^5,16,19,34^ These increased costs may have impacted patient medication costs and indirect costs to facilitate new medication acquisition. Patients ability to navigate the health care system to communicate with prescribers to replace a recalled medicine with an alternate has not been explored. A timely response is essential for medications taken daily for chronic conditions, but may be challenging when large number of persons are affected. Finally, the public’s trust in the health care system is eroding;^38^ having cascading reports of cancer-producing impurities reported across valsartan manufactures, other ARBs, and non-cardiac drugs (i.e., ranitidine, metformin) may have exacerbated this issue.^4,34,39,40^

## CONCLUSION

The national valsartan recall and subsequent shortage adversely impacted the outcomes of patients with hypertension. The downstream implications included increased hospitalization, emergency department, and urgent care encounters due to cardiac events. As the threat of shortages increases for drugs to treat common outpatient conditions, we must implement agile systems at the local- and national-levels to provide patients with therapeutic alternatives to prevent avoidable health care use and adverse outcomes.

## Data Availability

We are committed to collaborating and sharing these data to maximize their value to improve patients' health to the greatest degree possible. We can provide access to the programming code used to identify the sample and conduct analyses and will conduct additional analyses as requested. We cannot provide a link to the database directly, as the Optum Clinformatics Datamart dataset used for this study is proprietary and unable to be shared. Interested individuals may contact the corresponding author (Dr. KJ Suda) for information.

## Acknowledgements

The views expressed in this article are those of the authors and do not necessarily reflect the position or policy of the Agency for Healthcare Research and Quality, the Department of Veterans Affairs or the United States government.

